# Clinically Discordant Siblings with Spinal Muscular Atrophy: Insights from Patient-Specific iPSC-Derived Motor Neurons and Literature Review

**DOI:** 10.1101/2025.09.11.25335332

**Authors:** Sara L. Cook, Tyller Mensa, Henry Noma, Maya Jahnke, Noemi Vidal Folch, Christian Stout, Ron Hrstka, Sybil Hrstka, Lindsey Kirkeby, Devin Oglesbee, Linda Hasadsri, Duygu Selcen, Nathan P. Staff

## Abstract

Spinal muscular atrophy (SMA) is a neurodegenerative disease caused by homozygous loss of the *SMN1* gene. Copy number of the nearly identical paralog, *SMN2*, correlates with disease severity. *SMN2* is the primary modifier of SMA, with only a few other modifiers reported. We reviewed the literature of rare siblings affected by SMA that show discordance in clinical presentation despite having the same number of *SMN2* copies, which predicts the presence of genetic modifiers. We further recruited a sibling pair with discordant clinical presentations and performed detailed characterization. We utilized droplet digital PCR for deletion duplication testing and Sanger sequencing for full gene analysis of the *SMN2* gene which confirmed zero copies of *SMN1*, four copies of *SMN2*, and no *SMN2* modifying variants. Skin fibroblasts from each sibling were collected, reprogrammed into iPSCs, and differentiated to motor neurons. Patient-specific motor neurons revealed similar levels of SMN protein between the two siblings. Patient-specific iPSC-derived motor neurons collected from discordant siblings reported here may represent a powerful model for the discovery of SMN-independent modifiers.

## 1. Introduction

Spinal muscular atrophy (SMA) is an autosomal recessive neurodegenerative disease occurring in one out of every 10,000 live births [1, 2]. Patients with SMA exhibit degeneration of the alpha motor neurons of the spinal cord leading to progressive muscle weakness [3, 4]. SMA is a clinically heterogeneous disease, divided into five different types. Type 0 SMA is characterized by diminished fetal movements and subsequent presentation at birth with asphyxia and severe hypotonia [5]. Type 1 SMA presents before six months of age with hypotonia, abnormal breathing patterns, and progression to respiratory failure and death before two years of age [4]. Patients with Type 2 SMA present with a delay in gross motor milestones by 6–18 months of life, rarely stand and never walk, and suffer from progressive weakness, scoliosis, and restrictive lung disease [4]. Patients with Type 3 SMA achieve walking, but often require wheelchair assistance by adolescence [4]. The mildest type of SMA, Type 4, generally presents with mild proximal weakness with a later age of onset that progresses slowly [4].

SMA is caused by pathogenic variants, usually large deletions, in the survival motor neuron 1 (*SMN1*) gene which codes for the survival motor neuron (SMN) protein [6, 7]. The wide range of disease severity in SMA is largely due to the copy numbers of the paralog survival motor neuron 2 (*SMN2*), which differs from the *SMN1* gene by five nucleotides [6, 7], all of which are in non-coding regions, except for the cytosine to thymine variation in exon 7 [6, 7]. This variation is translationally silent; however, it causes alternative splicing such that 90% of transcripts are missing exon 7 and results in an unstable SMN protein that is rapidly degraded [8-12]. Accordingly, there is an inverse relationship between *SMN2* copy number and disease severity. In general, Type 0 SMA have one copy of *SMN2*, Type 1 SMA patients have two copies of *SMN2*, Type 2 SMA patients have three copies of *SMN2*, Type 3 SMA patients have three or four copies of *SMN2*, and Type 4 SMA patients have four or greater copies of *SMN2* [13, 14].

In addition to copy number variation, variants in the *SMN2* gene can modify disease severity. Variants c.859G>C and A-44G (c.835-44A>G) are known to associate with a mild SMA phenotype that does not correlate with *SMN2* copy number [15-17]. Other potential modifying variants, such as the A-549G (c.724-549A>G) and C-1897T (c.724-1897C>T) variants, have been associated with discordant phenotypes [18].

In the last decade, three disease-modifying therapies, nusinersen, onasemnogene abeparvovec, and risdiplam, have been developed for the treatment of SMA. These therapies are effective as they enhance SMN expression [19-21]. However, not all patients respond to these therapies, and many still suffer from severe impairments [19-21]. There is an urgent need to supplement current SMN-dependent therapies with potential SMN-independent therapeutic approaches.

Rarely, SMA families exhibit intrafamilial variation in severity of clinical presentation. In some SMA families, affected siblings have discordant clinical presentation despite having the same copy number of *SMN2* [22]. This suggests the presence of genetic modifiers, making discordant siblings an apt natural experimental model to investigate SMN-independent modifiers. Investigations of such families have led to the discovery of modifiers such as *PLS3, NCALD*, and *TLL2* [23-25]. Discordant siblings are rare and disappearing as clinical discordance will likely be masked by early treatment with disease-modifying therapies [22].

We have identified a sibling pair with discordant clinical presentation. We characterize the clinical discordance in this sibling pair, along with reviewing the literature on reported cases of discordant siblings. Furthermore, we characterize SMN levels in patient-specific induced pluripotent stem cell-derived motor neurons (iMNs) collected from the siblings.

## 2. Materials and Methods

### 2.1. Ethics Declaration

Results and available clinical information were acquired from patients included in this study; approval was granted by the Mayo Clinic Investigational Review Board (IRB # 19-004374) and informed consent from patients was obtained.

### 2.2. Review of Literature

A literature review was performed using PubMed® database using search terms “spinal muscular atrophy,” “discordant siblings,” and “intrafamilial variability.”

### 2.3. SMA Deletion/Duplication Analysis

The SMA deletion/duplication assay uses novel droplet digital PCR (ddPCR) technology to accurately quantify *SMN1* exon 7 NC_000005.9 (NM_022874.2) and *SMN2* exon 7 NC_000005.9 (NM_022876.2) by counting copies of the c.744C in *SMN1* and the c.744T in *SMN2* [26]. QuantaSoft Analysis Pro version 1.0.596 (Bio-Rad) was used for droplet-cluster classification and Poisson function application was used for the determination of absolute copy numbers [26].

### 2.4. *SMN2* Full Gene Analysis

The *SMN2* full gene sequencing assay utilizes long-range PCR of exons 2-8 of *SMN2* gene (NM_017411.4) followed by bidirectional Sanger sequencing to identify variants. Exon 1 of *SMN2* is PCR-amplified and bidirectionally Sanger-sequenced to identify variants in exon 1. Variants detected in *SMN2* exon 1 cannot be distinguished from variants in *SMN1* exon 1.

### 2.5. Reprogramming

2-4mm dermal punch biopsies were collected prospectively from eligible subjects. Tissue was placed in collection media and transported at room temperature. Tissue was subsequently rinsed in collection media and placed in a 10 cm dish with media. The biopsy was minced and selected pieces were transferred to a dish and covered with a coverslip and covered in media. The plate was incubated for four days, and media was changed every 5-7 days. Fibroblasts were reprogrammed using Cytotune-iPS 2.0. Karyotypically normal iPSC clones for each line that passed quality control testing for pluripotency. Quality control of iPSC lines included karyotyping (data not shown), immunofluorescence detection of pluripotency markers TRA-1-60 (R&D Biosystems, 1:100), Nanog (Cell Signaling, 1:100), SSEA4 (R&D Biosystems, 1:100), and Oct4 (Abcam, 1:200) (Supplementary Figure 1), flow cytometric detection of pluripotency markers (data not shown), and trilineage differentiation (data not shown). The lines used in this study were from the discordant sibling pair described in this report (Table 2, Supplementary Table 1). iPSCs were maintained on LDEV-free Geltrex in mTeSR™ 1 and passaged every 4 days.

### 2.6. Cell Line

Human iPSC cell line (WC023i-SMA-GM232) was purchased from WiCell®, 7-month-old male with homozygous loss of *SMN1* exon 7-8, and two copies of *SMN2*, confirmed by SMA deletion/duplication analysis (Supplementary Table 1). iPSCs were maintained on LDEV-free Geltrex in mTeSR™ 1 and passaged every 4 days.

### 2.7. Motor Neuron Differentiation

Motor neuron differentiation was performed following a well-established protocol [27]. hPSCs were plated Matrigel-coated plates. The next day, the medium was replaced with media: DMEM/F12, Neurobasal medium at 1:1, 0.5×N2, 0.5×B27, 0.1mM ascorbic acid, 1×Glutamax, 1×penicillin/streptomycin, 3μM CHIR99021, 2μM DMH-1, and 2μM SB431542. The media was changed every other day for 6 days. The cells were then dissociated with Dispase (1 mg/ml) and split at 1:6 with media: DMEM/F12, Neurobasal medium at 1:1, 0.5×N2, 0.5×B27, 0.1mM ascorbic acid, 1×Glutamax, 1×penicillin/streptomycin, 0.1μM RA and 0.5μM Purmorphamine, 1μM CHIR99021, 2μM DMH-1, and 2μM SB431542. The medium was changed every other day for 6 days. Next, the cells were expanded with media: DMEM/F12, Neurobasal medium at 1:1, 0.5×N2, 0.5×B27, 0.1mM ascorbic acid, 1×Glutamax, 1×penicillin/streptomycin, 3μM CHIR99021, 2μM DMH-1, 2μM SB431542, 0.1μM RA, 0.5μM Purmorphamine, and 0.5 mM VPA. They were split 1:6 once a week with Dispase (1 mg/ml). Next, the cells were dissociated with Dispase (1 mg/ml) and cultured in suspension in media: DMEM/F12, Neurobasal medium at 1:1, 0.5×N2, 0.5×B27, 0.1mM ascorbic acid, 1×Glutamax, 1×penicillin/streptomycin, 0.5μM RA, and 0.1μM Purmorphamine. The media was changed every other day for 6 days. The cells were then dissociated with Accumax into single cells and plated on Matrigel-coated plates. The cells were cultured with media: DMEM/F12, Neurobasal medium at 1:1, 0.5×N2, 0.5×B27, 0.1mM ascorbic acid, 1×Glutamax, 1×penicillin/streptomycin, 0.5μM RA, 0.1μM Purmorphamine, and 0.1μM Compound E for 10 days. iMN maturity was assessed by the detection of motor neuron-specific marker (*MNX1, ISL1*, and *CHAT*) transcription via quantitative reverse transcription PCR and by the detection of expression of motor neuron-specific markers (HB9, ISL-1, and ChAT) via immunofluorescence cell staining (Supplementary Figure 2).

### 2.8. Immunofluorescence

iMNs were fixed in 4.0% paraformaldehyde (Sigma Aldrich) solution in phosphate-buffered saline (PBS), permeabilized in 0.25% TritonX (Sigma Aldrich), and blocked in SuperBlock (PBS) Blocking Buffer (Thermo Scientific). Primary antibodies to ISL-1 (Abcam, 1:50), HB9 (Invitrogen, 1:50), ChAT (Proteintech, 1:100), and Tuj-1 (Proteintech, 1:1000) were added, and counter-stained with AlexaFluor secondary antibody (Chromotek, 1:2000). Nuclei were labeled with DAPI. Cells were imaged using Agilent BioTek Cytation and Zeis Axio Observer Z1.

### 2.9. Western Blot

Cells were lysed with RIPA lysis buffer (Protein Simple) containing HALT phosphatase inhibitor (100x, Thermo Scientific), HALT protease inhibitor (100x, Thermo Scientific), and 0.5 M EDTA (100x, Thermo Scientific) on ice for 30 minutes, vortexing every 5 minutes. The lysates were centrifuged 13,000g for 20 minutes at 4 degrees C. The lysates were measured with the Bradford protein assay (BioRad). Protein samples (10-25 μg) from each group were subjected to 10% polyacrylamide gel electrophoresis and transferred onto nitrocellulose membranes. After being blocked with EveryBlot blocking buffer (BioRad), the membranes were incubated with mouse anti-SMN (BD Transduction Laboratories, 1:250 to 1:5000 depending on the trial) and mouse anti-Tuj1 (ProteinTech, 1:5000 to 1:7500) at 4 °C overnight. The membranes were washed with TBST, incubated with anti-mouse horseradish peroxidase-conjugated secondary antibodies (1:20,000, Jackson ImmunoResearch Laboratories) at 20–25 °C for 1 h, and visualized via enhanced chemiluminescence by using the SuperSignal West Pico PLUS ECL detection kit (Thermo Fisher Scientific). Quantification was performed using ImageJ., and was performed on seven independent trials, detecting SMN levels with Tuj1 as a loading control with all samples normalized to Tuj1. Each trial contained two iPSC-derived clones for each sibling, levels were averaged together and normalized to the levels of the Type 1 SMA patient.

### 2.10. Statistical Analysis

Statistical analyses were completed using OriginLab (version 10.2). An ANOVA test was used to detect significant differences between groups. Tukey’s multiple comparison test was used to determine the significance of the conditions tested. A Student’s T-test was used to compare the means of two groups. P < 0.05 was considered statistically significant.

## 3. Results

### 3.1. Review of Reported Cases

We performed a literature review for reports of discordant siblings. Intrafamilial variability has been reported in the literature as early as 1967 [28]. Following this initial report, many cases of discordant families were reported; however, many early cases did not report *SMN2* copy numbers (Supplementary Table 3) [29-37]. We focused our literature review on cases with reported *SMN2* copy numbers.

In total, we identified 27 families with SMA discordant siblings having identical *SMN2* copy numbers (Table 1) [23, 24, 38-47]. Of these cases, 14 families consist of an asymptomatic sibling and a sibling with either Type 2 or Type 3 disease. 1 family consists of a sibling pair discordant with Type 1 vs Type 2 disease, a discrepant age of onset of 3 months. 4 families include a sibling pair discordant with Type 2 vs Type 3 disease, a discrepant age of onset of up to 11 years. 2 families include a sibling pair discordant with Type 3 vs Type 4 disease, a discrepant age of onset of up to 24 years. 6 families consist of a sibling pair discordant, both with Type 3 disease, a discrepant age of onset of up to 16 years.

**Table 1.**
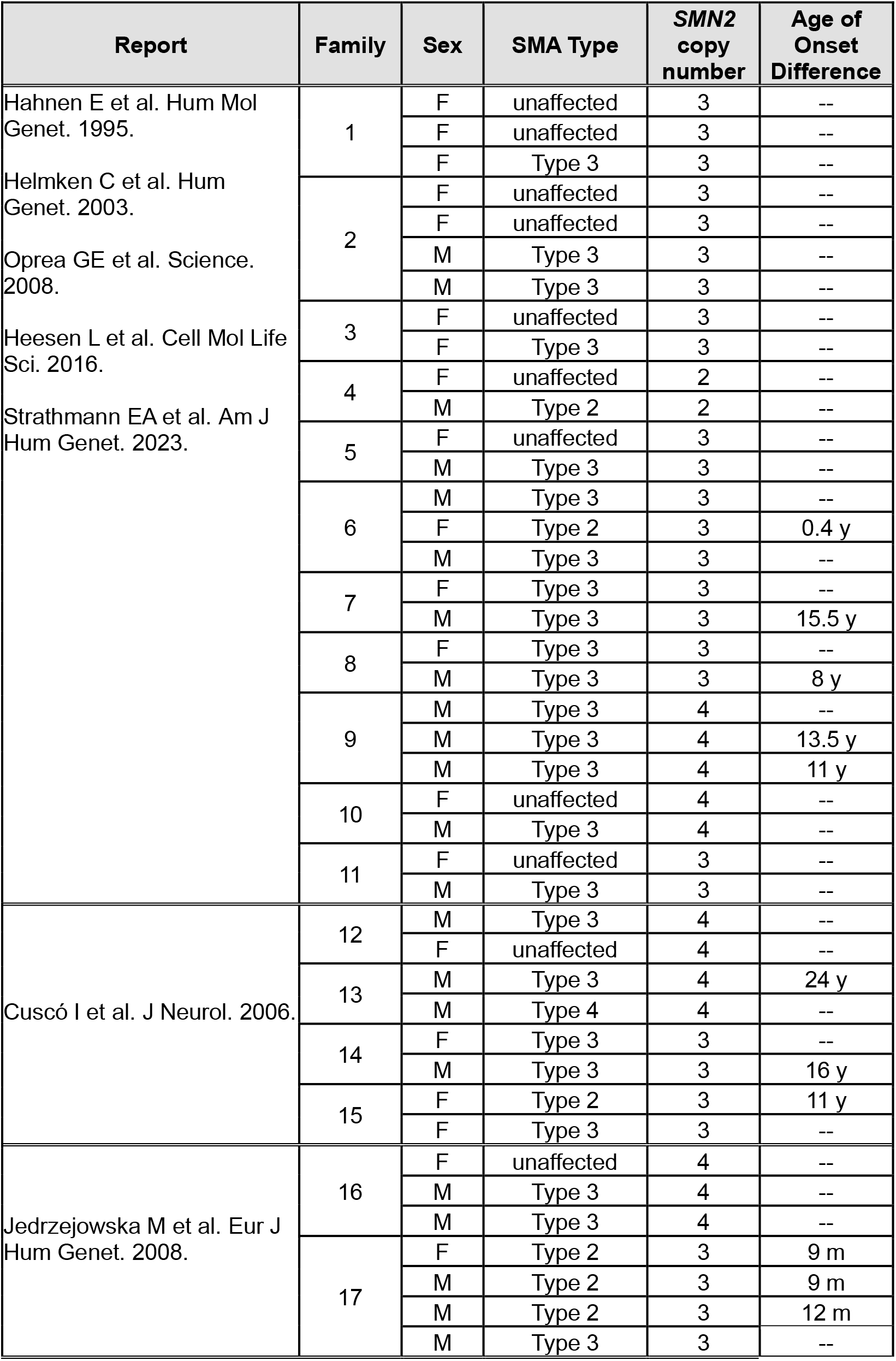
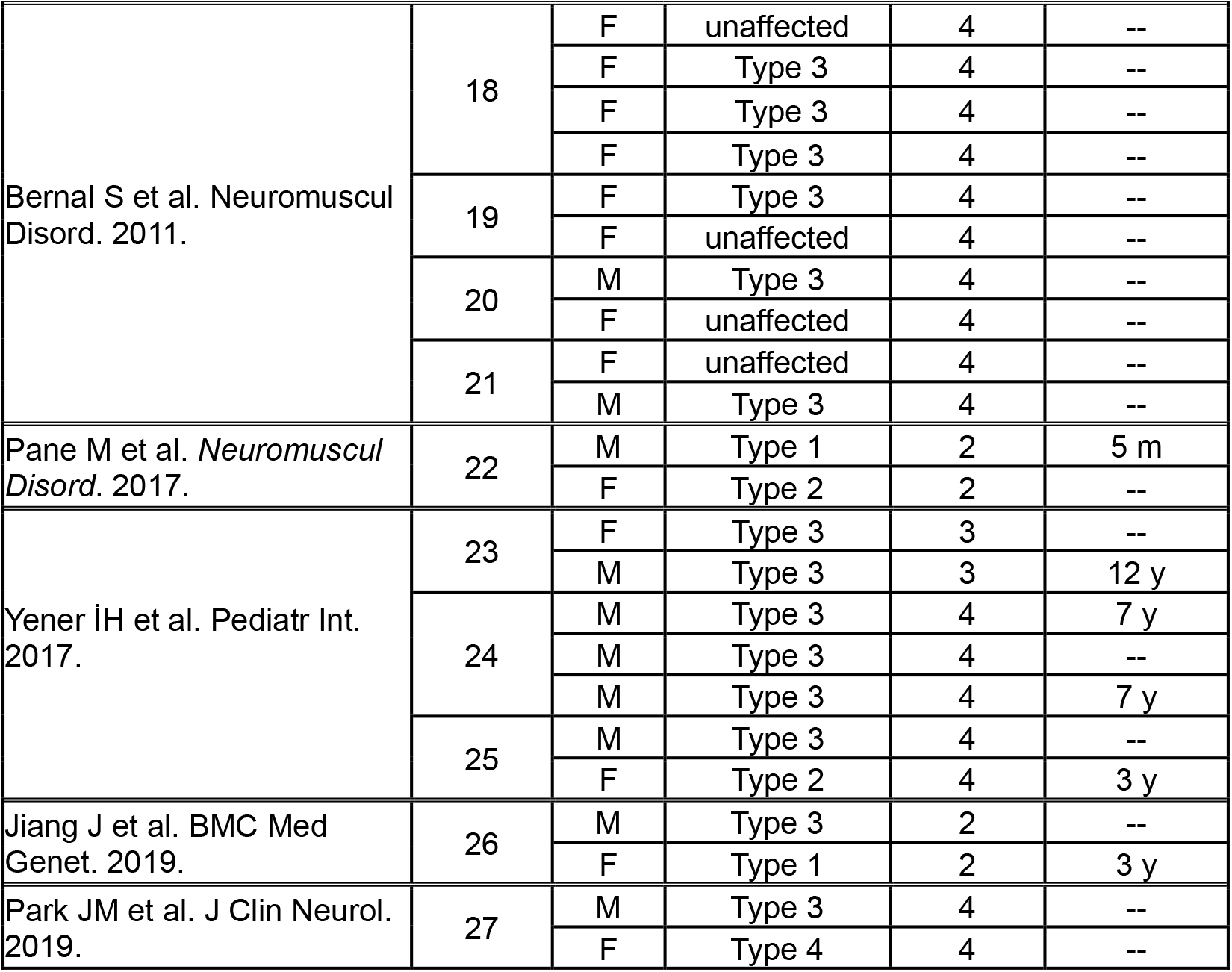
Reported Cases of Discordant Siblings

Most discordant families have high copy numbers of *SMN2* (Table 1). 12 families reported to have three copies of *SMN2*, and 12 families reported to have four copies of *SMN2*, while only 3 families reported to have two copies of *SMN2*. 18 of the 27 families consist of discordant siblings of different sexes, 16 of which reported the female sibling as the less affected sibling (Table 1).

### 3.2. Discordant Sibling Clinical Presentations

We identified two siblings affected by SMA with discordant clinical presentations. Patient 1 presented with a waddling gait and intermittent fine tremor in his hands.

He was monitored using the North Star Ambulatory Assessment, the Hammersmith Functional Motor Scale - Expanded for SMA (HFSME), the Revised Upper Limb Module for SMA (RULM), and the Six-Minute Walk Test (6MWT) (Figure 1). Over time his motor strength and motor assessment testing scores have shown improvement but remained below expected levels for his age (Figure 1).

**Figure 1.**
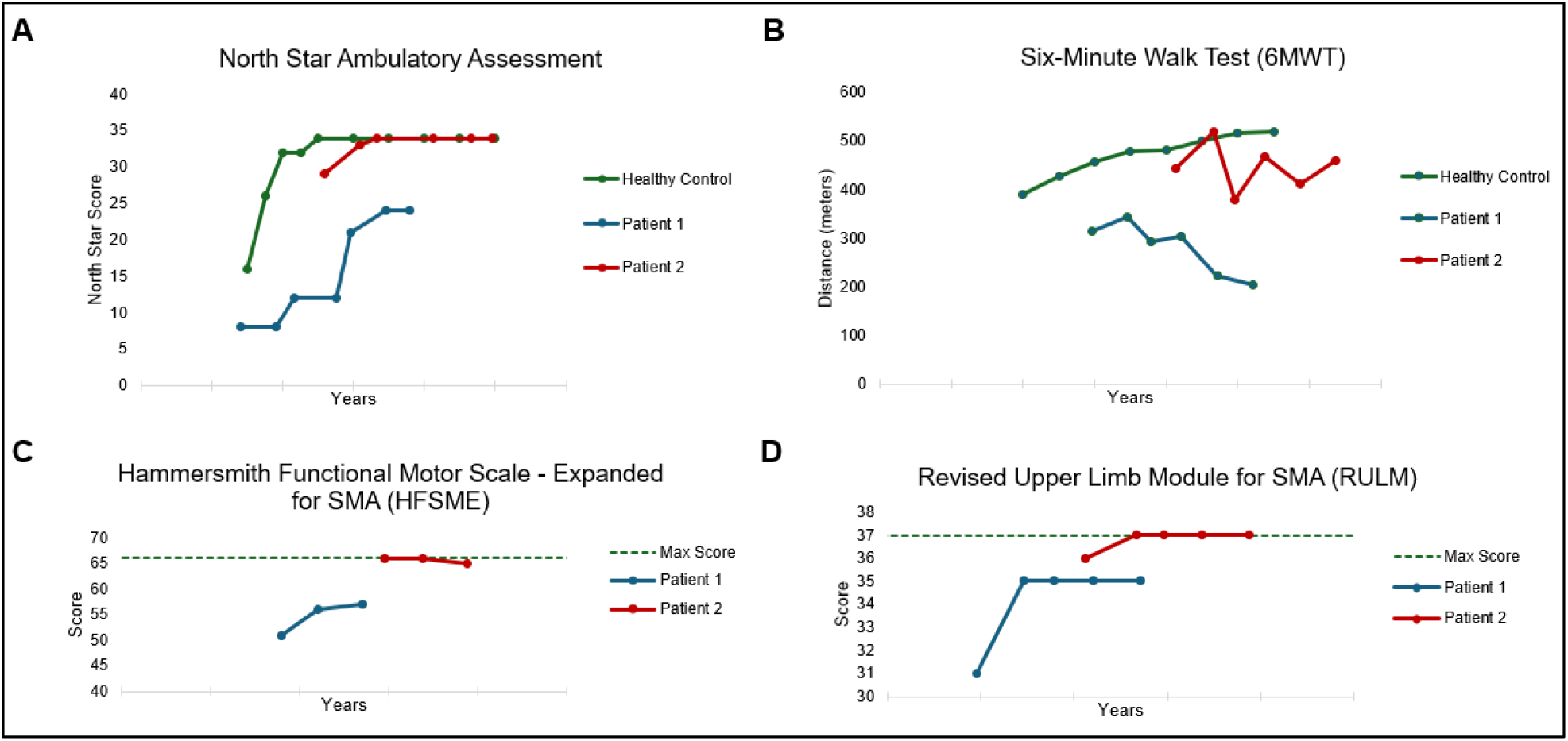
Motor Assessment Testing. (A) North Star Ambulatory Assessment of the patients over time. The healthy control represents the score achieved by at least 85% of healthy children at that age. (B) Six-Minute Walk Test of the patients over time. The healthy control represents the 50^th^ percentile distance traveled in 6 minutes in a population of healthy children at that age. (C) Hammersmith Functional Motor Scale-Expanded for SMA of the patients over time. The maximum score is 66. (D) Revised Upper Limb Module for SMA of the patients over time. The maximum score is 37. Tick marks on X-axis denote 2 years.

Patient 2 presented with subtle signs of weakness; frequent falls and ran with short strides. Her age of onset was over two years greater than the age of onset of Patient 1.

She was monitored using the same motor assessment testing as Patient 1 (Figure 1). Over time, her motor strength and motor assessment testing scores improved and met age-based normative values (Figure 1).

The siblings’ age of onset, clinical presentations, motor strength, and motor assessments reveal a discordance in their disease severity (Figure 1).

### 3.3. Molecular Characterization of *SMN1* and *SMN2*

Commercial genetic testing of both patients performed at an external diagnostic laboratory revealed homozygous deletion of *SMN1* and ≥4 copies of *SMN2*. Diagnostic testing was repeated at our internal reference laboratory to determine the exact copy number of *SMN2* (Figure 2a-b). ddPCR deletion/duplication analysis of both patients revealed an absence of *SMN1* positive clusters and absolute copy number analysis revealed that both patients have zero copies of *SMN1* and four copies of *SMN2* (Figure 2a-b).

To rule out modifying variants in the *SMN2* gene, *SMN2* full gene sequencing analysis was performed. Sanger sequencing detected the c.462A>G variant in exon 4 of both patients (Figure 2b-c). Both patients are heterozygous for this variant; the variant present in at least one copy of *SMN2*. This is not a conserved nucleotide (phyloP: 0.00 [-19.0, 10.9]) and this variant is not predicted to impact splicing; GeneSplicer predicted a 3’ splice site with a score of 3.8 and NNSPLICE predicted a 5’ splice site with a score of 1.0 and a 3’ splice site with a score of 0.97. [50-52]. This variant is synonymous (p.Gln154=) and therefore is not predicted to impact the gene product.

**Figure 2.**
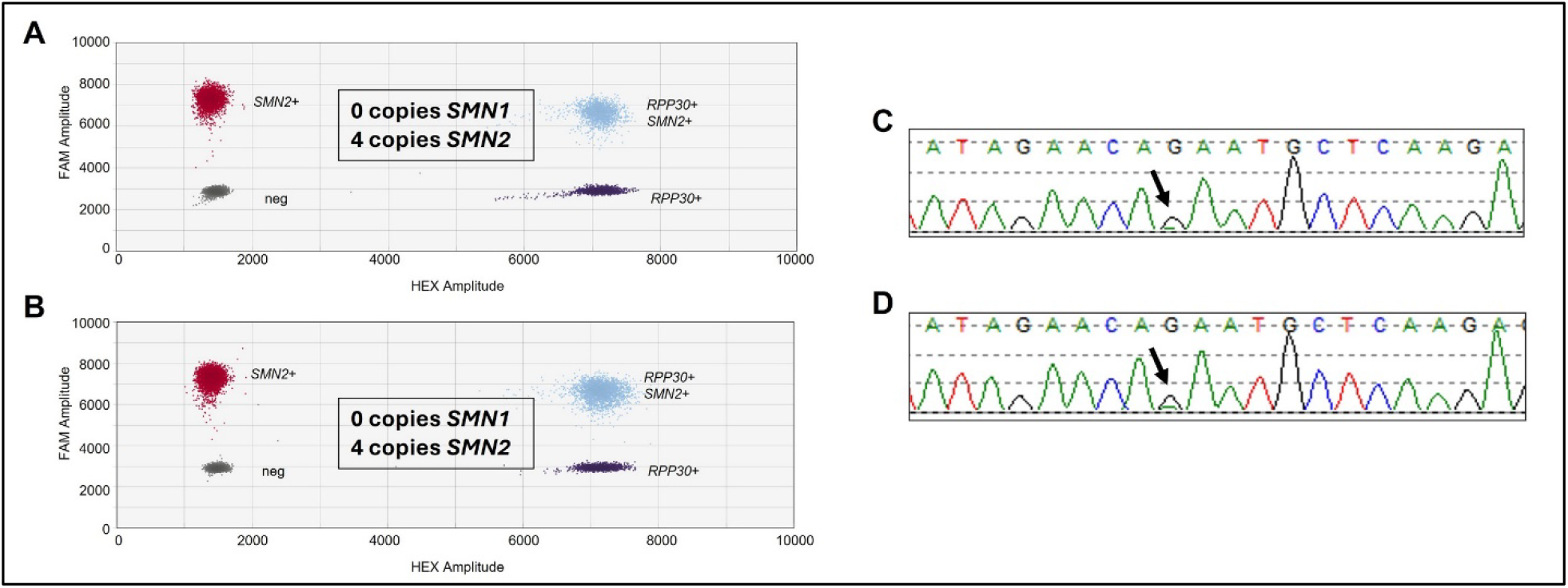
Molecular Genetic Testing. SMA deletion/duplication analysis via ddPCR from Patient 1 **(A)** and Patient 2 **(B)**. Droplet clusters correspond to populations of nucleotide targets; *SMN1, SMN2*, or *RPP30* (control), and resulting CNV of *SMN1* and *SMN2* for each sample. Sanger sequencing electropherogram of *SMN2* from the Patient 1 **(C)** and Patient 2 **(D)** Arrow notes c.462A>G, benign single nucleotide variant in exon 4, present in both patients.

This molecular characterization confirmed concordant copy numbers of *SMN2* and concordant full gene analysis of *SMN2* between the siblings, with no modifying variants in *SMN2* (Figure 2).

### 3.4. SMN Levels in Patient-Specific iMNs Collected from Discordant Siblings

Patient-specific iMNs from the siblings were utilized as a disease model to investigate levels of SMN. Skin fibroblasts from each sibling were collected and reprogrammed. iPSCs were positive for pluripotent markers Nanog, Oct4, TRA-1-60, and SSEA4 (Supplementary Figure 1). iPSCs were differentiated to motor neurons. Transcriptional profiling of fully differentiated iMNs revealed high expression of motor neuron-specific markers, *ISL1, MNX1*, and *CHAT* as compared to expression levels in iPSCs (Supplementary Figure 2a). Immunofluorescent cell staining of fully differentiated iMNs revealed positive staining for Tuj1 and motor neuron-specific markers, ChAT, ISL-1, and HB9 (Supplementary Figure 2b).

Western blot analysis was performed on the patient-specific iMNs collected from Patient 1 and Patient 2, along with iMNs collected from an SMA patient with Type 1 disease (two copies of *SMN2*) (Figure 3a). This analysis revealed both Patient 1 and Patient 2 to have higher levels of SMN compared to the Type 1 SMA patient (p<0.05) (Figure 3b). The SMN levels from Patient 1 and Patient 2 were equivalent (p=1.0) (Figure 3b).

**Figure 3.**
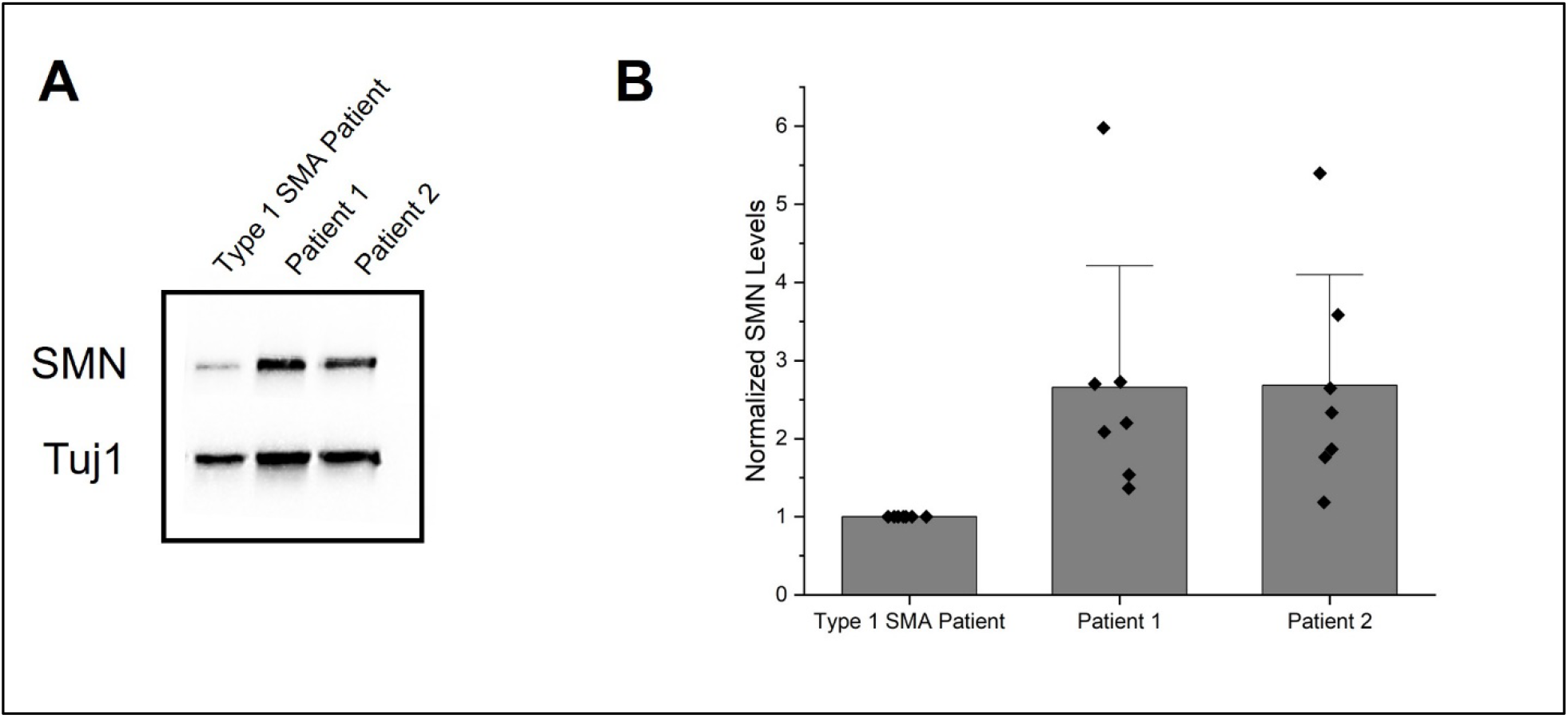
SMN Levels in Patient-Specific iPSC-Derived Motor Neurons. (A) Representative western blot of iPSC-derived MNs from a Type 1 SMA patient, Patient 1, and Patient 2, blotted for SMN and Tuj1 as a loading control. (B) Quantification of western analysis of 7 independent trials detecting SMN levels, Tuj1 as a loading control, each trial contained two iPSC-derived clones for each patient, clones averaged together per trial. A one-way ANOVA revealed a statistically significant difference in mean SMN levels [F(2, 18) = (4.41), *p*-value < 0.05]. Tukey’s HSD Test for comparisons found that the mean SMN levels were significantly different between the Type 1 SMA patient and the patients (*p*<0.05), and no statistically significant difference in mean SMN levels between Patient 1 and Patient 2 (the discordant sibling pair) (*p*=1.0).

## 4. Discussion

We identified two siblings affected by SMA with discordant clinical presentations. The male patient is the more affected sibling, having an age of onset 2 years less than his female sibling.

An association between sex and disease severity in discordant sibling cases was also observed in our literature review. Our review of reported cases of discordant siblings revealed that the majority of reported cases are of different sexes, and of these cases with different sexes, the majority of the less affected siblings are female (Table 1). Our literature review was limited to reports with specific relevant search terms and therefore may not have included all cases that have been reported.

Sex-specific phenotypic variability has been reported in SMA. The SMA TREAT-NMD dataset revealed that more male family members are affected by SMA than female family members [53]. However, in a study investigating 303 SMA sibships using the Cure SMA membership database, there was no association between sibling sex and concordance [22]. Of note, the data collected in this database was obtained per patient report and *SMN2* copy numbers are not reported. Furthermore, in a study analyzing data collected from both the TREAT-NMD Global SMA Registry and the Cure SMA membership database, it was reported that symptoms were more severe in males than females in Types 2 and 3 SMA, and that motor function scores were higher in females than males in Types 1 and 3 SMA [53]. Of the few known SMA disease modifiers, multiple modifiers are encoded by genes on the X chromosome, and higher expression of some of these modifiers has been associated with milder phenotypes in female patients [24, 54-56].

The siblings we report here have discordance in disease severity. As such, we have confirmed they have concordant molecular testing. Both siblings have four copies of *SMN2* (Figure 2a). Full gene sequencing of *SMN2* revealed both siblings to have a single nucleotide variant in exon 4, c.462A>G (Figure 2b). This variant has been reported in *SMN2* [14, 18]. In a large study of 217 SMA patients the c.462A>G allele was detected frequently and was not found to correlate with patients with clinical presentations that were more mild or severe than expected [18].

Our analysis only included exons 1-7. Therefore, exon 8, which codes for the UTR, and the intronic regions were not sequenced. There are known modifying variants in intronic regions, including the A-44G (c.835-44A>G) variant in intron 6 [17]. Therefore, we cannot rule out differential variants in the siblings that may be present in these regions. Future studies include long-read sequencing of the SMN locus to rule out such variants. Long-read HiFi sequencing approaches have been shown to successfully determine copy number variation, detect phased variants, and identify *SMN1* and *SMN2* single-nucleotide variants, small insertions, and deletions [57, 58].

Following the concordant molecular characterization, SMN levels from iMNs were analyzed. SMN protein levels were not different between the sibling-specific iMNs (Figure 2b). Determining precise levels of SMN that correlate with phenotype is difficult, and detecting minor differences in expression levels of SMN is technically challenging. Moreover, we did not investigate levels of SMN in earlier progenitor stages of differentiation and can therefore not rule out differential expression levels at earlier time points. Future studies include using additional platforms for quantitative SMN detection, such as an electrochemiluminescence (ECL)-based immunoassay, which have been reported to have a lower limit of SMN detection of 3 pg/mL [59], and to investigate SMN levels in earlier progenitor stages.

Furthermore, an in vitro iMN model may not replicate SMN levels in motor neurons in vivo. SMN expression may differ in the siblings at different developmental stages. High levels of SMN are detected in the spinal cord throughout development and are detected as early as 8 weeks [60]. Moreover, spinal cord SMN protein levels are higher in fetal samples than in postnatal samples [61].

Nonetheless, the molecular characterization of the siblings and SMN protein levels detected in patient-specific iMNs collected from the discordant siblings, collectively suggests that the observed phenotypic differences are potentially due to SMN-independent modifiers. As such, the discordant siblings in this study are a model for the discovery of SMN-independent modifiers. Further studies, which include transcriptomic and proteomic analysis, are needed to investigate potential modifiers.

Identifying SMN-independent genetic modifiers may uncover potential therapeutic targets. Modifiers previously discovered by investigating discordant siblings have proven to be potential therapeutic targets. The therapeutic potential of modulating PLS3 levels, an SMA modifier discovered by investigating SMA discordant siblings, has been investigated. SMN antisense oligonucleotide treatment in combination with PLS3 overexpression rescues survival and motor function in a severe SMA mouse model [56].

TLL2, another SMA modifier discovered by investigating discordant siblings, is an activator of myostatin, which inhibits skeletal muscle growth. Whole exome sequencing of discordant siblings revealed TLL2 variants in the milder sibling in that study [23]. The variants are predicted to reduce myostatin activation, which supports increased muscle growth [23]. Myostatin inhibitors have shown therapeutic benefit in mouse models for SMA and are currently in clinical trials to treat SMA patients [62, 63].

Discordant siblings, as described in this study, are a model for discovering SMN-independent modifiers. Disease-modifying therapies enhance SMN expression; however not all patients respond to these therapies, and many still suffer from impairments [19-21]. There is an urgent need to supplement current SMN-dependent therapies with SMN-independent therapeutic approaches. Further investigation for the discovery of SMN-dependent modifiers using patient-specific iMNs collected from the discordant siblings reported here holds significant potential for identifying new therapeutic targets and inciting new therapeutic strategies.

## Data Availability

The data produced in the present study are not readily available because of ethical and privacy restrictions. Requests to access the data should be directed to the corresponding author.

## 5. Acknowledgments

We would like to thank the Mayo Clinic Center for Regenerative Biotherapeutics Biotrust, Director Zachary Resch, PhD, Cass Gray, and Douglas Smith IV, as some of the data in this publication were produced in the Biotrust laboratory.

The authors thank the patients and their families for their participation in this study.

