## Supplementary Materials for "Clinically Discordant Siblings with Spinal Muscular Atrophy: Insights from Patient-Specific iPSC-Derived Motor Neurons and Literature Review"

**Supplementary Table 1.** *SMN1* and *SMN2* CNV Analysis of iPSC Lines.

| Patient | Line | *SMN1* CNV | *SMN2* CNV |
|---|---|---|---|
| Type 1 SMA Patient | 12BT16 232 | 0 | 2 |
| Patient 1 | 24BT5 46 | 0 | 4 |
| | 24BT5 105 | 0 | 4 |
| Patient 2 | 24BT23 68 | 0 | 4 |
| | 24BT23 71 | 0 | 4 |

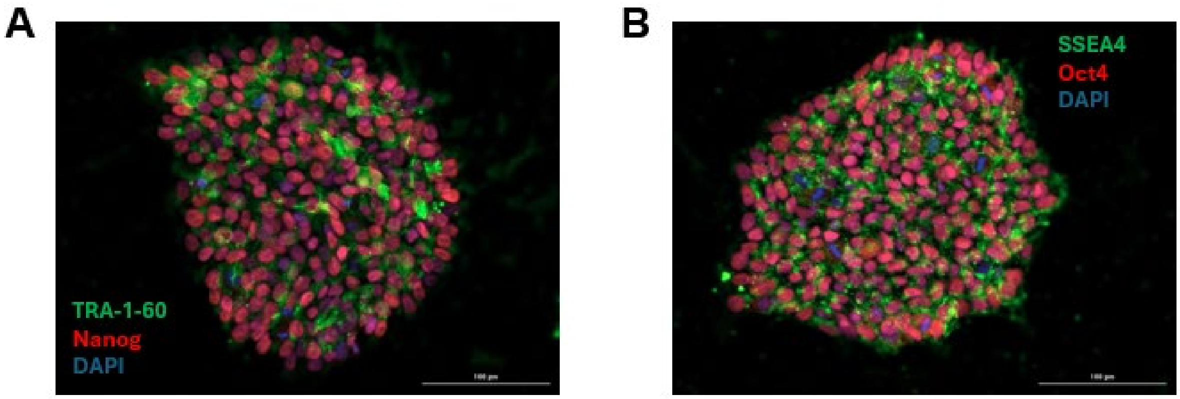


**Supplementary Figure 1: Quality Control of iPSC.** Clone-specific iPS cell colony (Passage < 10) stained for the pluripotency markers **(A)** Oct4 and SSEA, **(B)** Nanog and TRA-1-60 with a nuclear counterstain (DAPI). 20X magnification. Scale bar is 100um.

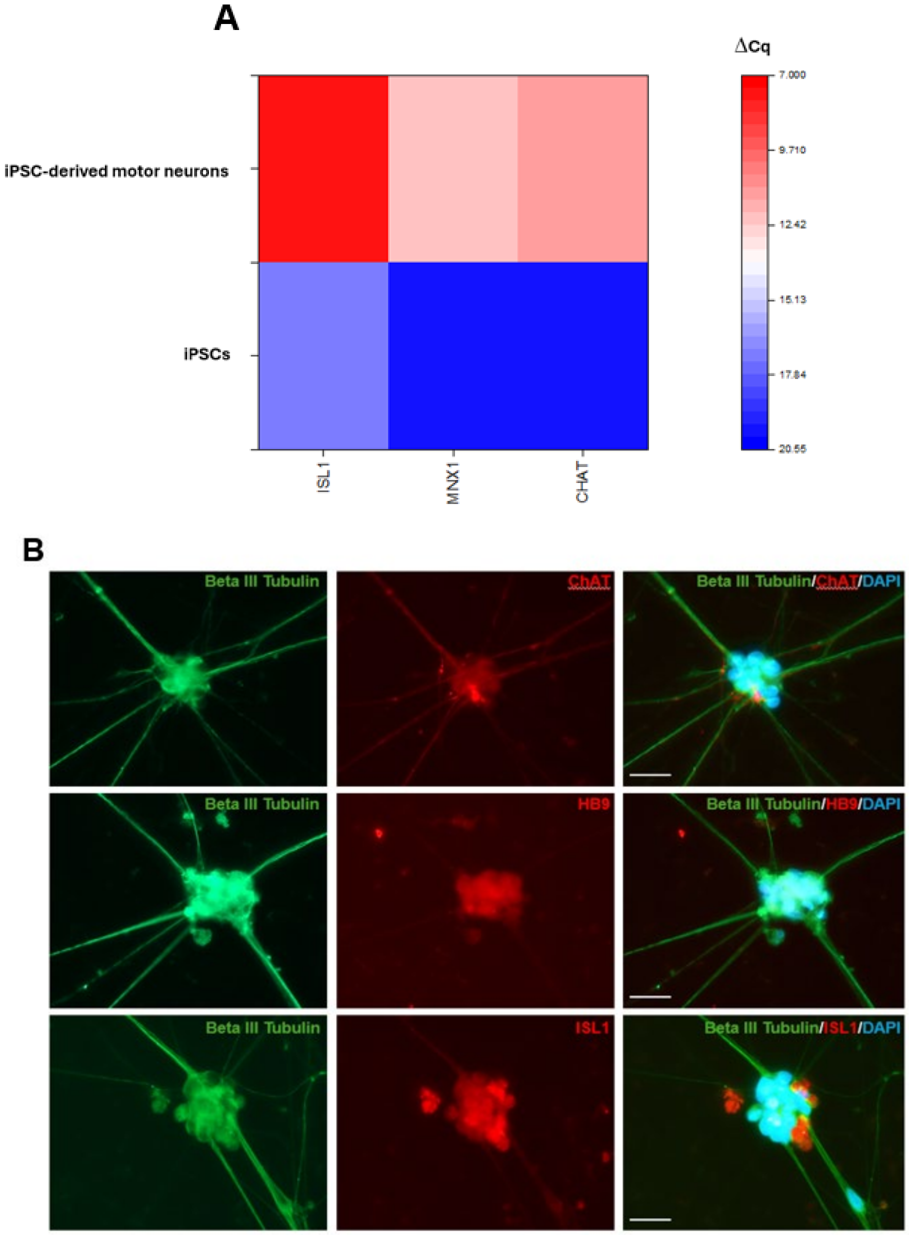


**Supplementary Figure 2: Transcriptional Profiling and Immunofluorescence Detection of Motor Neuron-Specific Markers of iPSC-Derived Motor Neurons. (A)** Heatmap of RT-PCR ΔCq values for *ISL1*, *MNX1*, and *CHAT* from representative iPSC-derived motor neurons and iPSCs. Three replicates were performed, and a Student's T-test was used to compare the means of the iPSC-derived motor neurons and iPSCs. ISL1: $t(4) = 20.78546$, $p < 0.0001$, MNX1: $t(4) = 11.2492$, $p < 0.0005$, CHAT: $t(4) = 18.48626$, $p < 0.0001$. **(B)** Representative iPSC-derived MNs were stained for motor neuron-specific markers ChAT, HB9, Isl1, and with neuron-specific beta III tubulin and with a nuclear counterstain (DAPI). 40X magnification. Scale bar is 20um.

**Supplementary Table 2.** Primer Sequences.

| Gene | Forward | Reverse | Product Size (bp) | Tm (°C) |
|---|---|---|---|---|
| **ISL1** | 5'-GTGTGATCCGGGTCTGGT-3' | 5'-GCGAAGTCGCTCAGTACTTTC-3' | 226 | 57.0, 55.8 |
| **MNX1** | 5'-CTAAGATGCCCGACTTCAACT-3' | 5'-CCGGTTCTGGAACCAAATCT-3' | 206 | 55.0, 54.5 |
| **CHAT** | 5'-TATGGGCTCTTCTCCTCCTAC-3' | 5'-CACTGAGACGGCGGAAATTA-3' | 157 | 55.1, 54.9 |
| **β-ACTIN** | 5'-TCCACGAAACTACCTTCAACTC-3' | 5'-AGGGCAGTGATCTCCTTCT-3' | 139 | 54.5, 55.3 |
| **TUBB3** | 5'-CAAGATGTCCTCCACCTTCATC-3' | 5'-GACACCAGGTCGTTCATCTT-3' | 177 | 55.2, 55.1 |

**Supplementary Table 3.** Reports of SMA Discordant Siblings, *SMN2* CNV Not Reported.

| Report | Family | Sex | Clinical Presentation |
|---|---|---|---|
| Prot J et al. Neurol Neurochir Pol. 1969. | 1 | M | Type 2 |
| | | F | Type 3 |
| | | F | Asymptomatic |
| | 2 | F | Type 2 |
| | | F | Type 2 |
| | | F | Type 2 |
| | | F | Asymptomatic |
| | | M | Asymptomatic |
| | 3 | M | Type 3 |
| | | M | Asymptomatic |
| | 4 | M | Type 2 |
| | | M | Asymptomatic |
| | | M | Asymptomatic |
| | 5 | F | Type 3 |
| | | M | Asymptomatic |
| Müller B et al. Am J Hum Genet. 1992. | 6 | F | Type 3 |
| | | M | Type 2 |
| | 7 | M | Type 2 |
| | | F | Type 3 |
| | 8 | F | Type 3 |
| | | M | Type 2 |
| | 9 | M | Type 2 |
| | | F | Type 3 |
| | | F | Type 3 |
| Brahe C et al. Am J Med Genet. 1993. | 10 | M | Type 3 |
| | | M | Type 3 |
| Cobben JM et al. Neuromuscul Disord. 1993. | 11 | M | Asymptomatic |
| | | M | Type 3 |
| | | F | Type 3 |
| Burghes AH et al. Hum Genet. 1994. | 12 | F | Type 2 |
| | | F | Type 3 |
| | 13 | F | Type 2 |
| | | F | Type 2 |
| | | M | Type 3 |
| | 14 | F | Asymptomatic |
| | | M | Type 3 |
| Rudnik-Schöneborn S et al. Am J Med Genet. 1994. | 15 | F | Type 1 |
| | | F | Type 1 |
| | 16 | M | Type 1 |
| | | F | Type 1 |
| | 17 | M | Type 1 |
| | | F | Type 1 |
| | | M | Type 1 |
| | 18 | F | Type 1 |
| | | F | Type 2 |

| | | | |
|---|---|---|---|
| | 19 | M | Type 1 |
| | | F | Type 2 |
| | 20 | F | Type 1 |
| | | M | Type 2 |
| | 21 | M | Type 1 |
| | | M | Type 2 |
| | 22 | M | Type 1 |
| | | M | Type 2 |
| | 23 | M | Type 2 |
| | | F | Type 2 |
| | 24 | M | Type 2 |
| | | F | Type 2 |
| | 25 | F | Type 2 |
| | | M | Type 3 |
| | | M | Type 3 |
| | 26 | M | Type 1 |
| | | M | Type 3 |
| | 27 | M | Type 3 |
| | | M | Type 3 |
| | 28 | M | Type 3 |
| | | M | Type 3 |
| | | M | Type 3 |
| | 29 | M | Type 3 |
| | | M | Type 3 |
| | | M | Type 3 |
| | 30 | F | Type 3 |
| | | F | Type 3 |
| | 31 | F | Type 3 |
| | | F | Type 3 |
| | 32 | M | Type 3 |
| | | F | Type 3 |
| | 33 | M | Type 3 |
| | | F | Type 3 |
| | 34 | F | Type 4 |
| | | F | Type 4 |
| | | M | Type 4 |
| Cobben JM et al. Am J Hum Genet. 1995. | 35 | F | Asymptomatic |
| | | F | Asymptomatic |
| | | M | Type 3 |
| | | M | Asymptomatic |
| | | M | Type 3 |
| Capon F et al. Neuromuscul Disord. 1996. | 36 | F | Asymptomatic |
| | | M | Type 3 |
| | 37 | M | Type 3 |
| | | F | Type 3 |
| McAndrew PE et al. Am J Hum Genet. 1997. | 38 | F | Type 2 |
| | | M | Asymptomatic |
